# Primary care during the COVID-19 pandemic – observations and challenges identified through a survey and interviews with community paediatricians: the calm before the storm

**DOI:** 10.1101/2020.12.30.20249037

**Authors:** Malte Kohns Vasconcelos, Katharina Weil, Daniela Vesterling-Hörner, Mehrsad Klemm, Tarik el Scheich, Hanna Renk, Katharina Remke, Hans Martin Bosse

## Abstract

**Background:** Globally, the COVID-19 pandemic has a major impact on healthcare provision. The effects in primary care are understudied.

**Aim:** To document changes in consultation numbers and patient management during the COVID-19 pandemic, and to identify challenges for patient care.

**Methods:** Survey of 44 paediatric primary care practices on consultation numbers and patient management changes (response rate 50%), and semi-structured interviews to identify challenges for patient care.

**Results:** Numbers of consultations for scheduled developmental examinations remained unchanged compared to the previous year while emergency visits were strongly reduced (mean 87.3 less/week in March–May 2020 compared to 2019, median reduction 55.0%). Children dependent on developmental therapy and with chronic health conditions were identified as patient groups receiving deteriorated care. High patient numbers, including of mildly symptomatic children presenting for health certificates, in combination with increased organisational demands and expected staff outages are priority concerns for the winter.

**Conclusion:** Primary care paediatricians offered stable service through the early pandemic but expect strained resources for the upcoming winter. Unambiguous guidance on which children should present to primary care and who should be tested would help to allocate resources appropriately, and this guidance needs to consider age group specific issues including high prevalence of respiratory symptoms, dependency on carers and high contact rates.

## Abstract (local language – German)

**Hintergrund:** Weltweit hat die COVID-19-Pandemie einen starken Einfluss auf die Gesundheitsversorgung. Die Effekte auf die Primärversorgung sind nicht ausreichend untersucht.

**Ziel:** Veränderungen in Konsultationszahlen und Patientenmanagement während der COVID-19-Pandemie beschreiben und Herausforderungen für die Patientenversorgung zu identifieren.

**Methoden:** Fragebogenstudie mit 44 Praxen aus der pädiatrischen Primärversorgung zu Konsultationszahlen und Änderungen im Patientenmanagement (Beantwortungsquote 50%) und halbstrukturierte Befragungen zur Identifikation von Herausforderungen für die Patientenversorgung.

**Ergebnisse:** Im Vergleich zum Vorjahr blieben Konsultationszahlen für geplante Vorsorgeuntersuchungen unverändert, während Notfallkonsultationen deutlich reduziert waren (Mittelwert 87.3 weniger pro Woche zwischen März und Mai 2020 im Vergleich zu 2019, Verringerung im Median um 55.0%). Kinder, die auf Entwicklungsförderung angewiesen sind, und Kinder mit chronischen Erkrankungen wurden als schlechter versorgte Patientengruppen identifiziert. Hohe Patientenzahlen, auch von Kindern mit milden Beschwerden, die für Gesundheitsbescheinigungen vorgestellt werden, in Kombination mit erhöhten organisatorischen Anforderungen und erwartete Ausfälle von Mitarbeitern sind wichtigste Sorgen für den Winter.

**Schlussfolgerung:** Niedergelassene Kinder- und Jugendärzte stellten in der frühen Pandemie eine stabile Versorgung sicher, erwarten aber eine Ressourcenknappheit für den kommenden Winter. Eindeutige Leitlinien dazu, welche Kinder in der Primärversorgung vorgestellt werden und welche getestet werden sollen, würden bei der geeigneten Ressourcenzuteilung helfen. Diese Leitlinien müssen altersgruppenspezifische Besonderheiten wie hohe Prävalenz von Atemwegssymptomen, Abhängigkeit von Versorgungspersonen und hohe Kontaktraten berücksichtigen.

## Introduction

Crises have been shown to result in decreased healthcare utilisation and increased complications from chronic health conditions like diabetes [21,23]. Studies on indirect health effects from infectious disease epidemics have focused on low-resource settings and have shown that disruption of routine healthcare likely resulted in similar mortality as the infectious disease itself and significant additional morbidity from chronic conditions [2,19]. Although effects from changed healthcare seeking behaviour are likely to be less pronounced in high-resource settings, concerns about missed regular follow-up visits and an expected increase in complications for children with chronic health conditions have also been voiced [6].

In the context of COVID-19, multiple studies have shown that unscheduled patient presentations were reduced during the first wave of the pandemic in hospital emergency departments [22], but it is unclear if non-hospital providers were partially compensating for this.

The aim of this study was to describe changes observed in paediatric primary care during the first wave of the COVID-19 pandemic and to identify challenges for the upcoming winter season.

## Method

The study consisted of a survey sent to paediatric practices in the academic teaching network and a subsequent set of interviews with paediatricians working in these practices. The practices were located in the Düsseldorf area in Western Germany that was among the first regions in Europe seeing high COVID-19 case numbers [26]. Primary care in Germany is provided through practices run by self-employed specialist physicians that are contracted to offer services to patients under public health insurance which is compulsory to the majority of the population. Under the terms of public insurance, paediatric specialists can only treat patients until their 18^th^ birthday.

### Survey

Between 26^th^ June and 3^rd^ July, 44 paediatric primary care practices in the Düsseldorf area in Western Germany were asked to complete a survey on patient management strategies and consultation numbers during the months of February through May. Survey participants were asked to estimate total consultation numbers during the survey period compared to previous years. Precise consultation numbers were collected separately for scheduled standard developmental assessments (German compulsory U and J examinations) and for unscheduled visits for one sample week per month (without a public holiday) and a respective, public holiday-free week from 2019. The method for obtaining these precise numbers was not specified. The survey form is provided in the online supplementary material for this article (document in German).

After entry of results into Microsoft Excel, data were imported into Stata 14, which was used for all further data management and statistical operations. Descriptive statistics were used and comparisons of consultation numbers between 2019 and 2020 were made by paired t-tests.

### Interviews

Care providers who responded to the survey and indicated their agreement to be contacted for an interview participated in semi-structured telephone interviews between 28^th^ July and 7^th^ August. The interviews were held and recorded via an online meeting tool (GoToMeeting by LogMeIn, Boston, MA). They consisted of 3 lead questions, structured short follow-up questions and open dialogue for clarification or further detail and were concluded with an open question for additional comments. The semi-structured interview lead questions were:

1. Which patients, in your opinion, receive better or worse paediatric outpatient care in the context of the current COVID-19 pandemic? Why? Are there solutions or ways to improve this situation?
2. What is your biggest worry for your practice or your caring for patients with regard to the upcoming winter? What could be done to address this worry?
3. Do you feel you are supplied with enough information on new findings on COVID-19 and the course of the pandemic to be able to care for your patients in the best possible way? What could be improved?

Audio recordings were transcribed, and transcripts were independently reviewed by two investigators. Responses were thematically grouped into clusters. Cluster disagreements between the investigators were resolved by discussion and, where no consensus was achieved, clustering was decided by majority vote among the co-authors. Per lead question, all clusters mentioned by at least two interviewees were listed.

### Ethical considerations

The study was approved by the ethics committee of the Medical Faculty of Heinrich Heine University Düsseldorf (study number: 2020-1056). All participants gave informed consent separately for participation in the survey and the interviews.

## Results

22 practices completed the survey (response rate of 50%). 10 of the practices were located in the urban centre of Düsseldorf and the remainder in the outskirts or surrounding communities. 19 practices (86%) had adopted a standardised case definition for COVID-19 suspect cases. Practices either adopted the case definition issued by the federal public health authority (Robert Koch Institute, RKI) directly or followed related case definitions issued by local public health authorities. Criteria for testing children for SARS-CoV-2 differed, with 59% reporting that they followed the definition of a suspect case for this decision (36% reported to follow this definition strictly and 23% reported making exceptions). 36% decided on a case by case basis without pre-specified criteria. 12 practices were able to supply numbers of SARS-CoV-2 tests performed in the specified weeks in February to May. As expected, numbers of patients tested for SARS-CoV-2 increased slightly over the study period with a maximum of 30 tests per week performed in one practice. Still, the median of tests per practice was low (0 in February, 1.5 in March and April and 2 in May). Only 3 practices could expect test results back on the same day, 13 (59%) received test results on the following day and 6 (27%) regularly had to wait for more than one day for test results to be back. Almost two thirds (64%) had already used serological testing and the majority (78%) of these reported deciding to do serology on a case-by-case basis.

All practices reported having changed clinic procedures in response to the pandemic. Most (91%) either used separate clinic hours for non-infection-related and infection-related consultations, used separate rooms for these groups or made both spatial and clinic hour changes (27% separated patients spatially, 27% temporally and 36% used both options). 11 practices (50%) had a policy to divert potentially SARS-CoV-2 infected patients to other facilities (e.g. hospital emergency departments or testing centres), either exclusively or in combination with patient separation. All but one of the practices estimated that their consultation numbers were lower than usual during the sampled period (mean 28% lower, i.e. 72%, 95% confidence interval 67% -- 77%). Figure 1a shows relative consultation numbers in March, April and May 2020 compared to corresponding weeks in 2019. Documented consultation numbers for developmental assessments were similar between 2019 and 2020, while emergency consultation numbers where markedly lower. Figure 1b shows the respective absolute numbers of consultations per week.

**Fig. 1:**
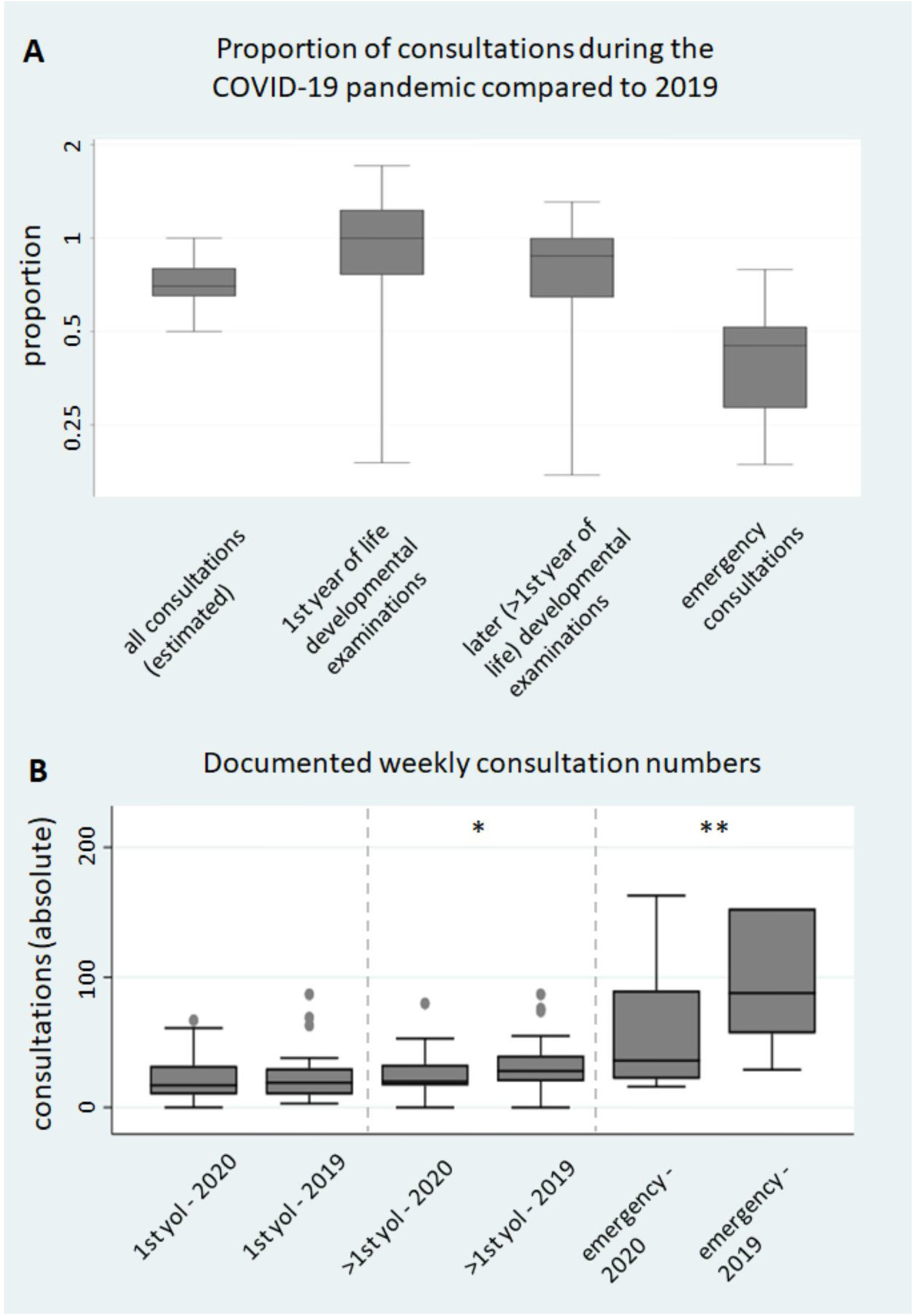
Patient consultation numbers in 2019 and 2020. Panel A: estimated proportion of patient consultations during the first pandemic wave as compared to regular numbers of consultation (left box), other boxes proportion of documented visits in sample weeks in March, April and May 2020 compared to corresponding weeks in 2019; 1st year of life (yol) developmental examinations comprise U1 – U6 examinations, later developmental examinations U7 – J2 examinations that are compulsory for all German children; Panel B: absolute weekly consultation numbers for sample weeks between March and May, 3 upper range outliers for right box (emergency – 2019) not shown; 1st yol and >1st yol indicating developmental examinations as specified for panel A; no difference for 1st yol (p=0.611), mean difference >1st yol 5.8 consultations per week (* p=0.002), mean difference emergency 87.3 consultations per week (** p=0.001)

All practices reported that parents had cancelled scheduled appointments due to the pandemic. 50% of practices said that they had cancelled and rescheduled patient appointments, either of their own initiative (82%) or following external regulations (18%). 64% of the practices that had cancelled appointments and 50% of the ones that had not cancelled appointments used telephone appointments as an alternative.

The majority of surveyed primary care providers reported that in their practice, patients were neither treated worse nor better during the pandemic (64% reported no patients were treated worse, 82% reported none were treated better). During the semi-structured interviews, 5 out of 8 interviewees reported that they observed that some patient groups generally received worse care during the pandemic. Table 1 lists patient groups that were named during the interviews.

**Table 1:** Patient groups identified to be perceived as receiving better or worse care during the COVID-19 pandemic and respective reasons

| **Better care during pandemic** |  |
| --- | --- |
| *Patient group* | *Advantages* |
| presenting for minor problems | fewer time constraints, easier access to the doctor |
| presenting for regular check-up visits | fewer time constraints |
| **Worse care during pandemic** |  |
| *Patient group* | *Issues* |
| chronic health conditions, incl. asthma and allergies | afraid to seek medical care or attend check-up visits, delayed adaptation of long-term medication |
| children with neurological and developmental problems | physiotherapy, occupational therapy and speech and language therapy suspended for extended periods of time |
| children from disadvantaged backgrounds, social paediatrics | suspended therapies, suspended social care visits; less reliably attending appointments |
| children requiring further investigations offered in hospitals | appointments unavailable |
| families with high fear of SARS-CoV-2 infection | afraid to seek medical care even if urgent |

The patient group most often named for receiving worse care was children in need of developmental and psycho-social support. In Germany this group is cared for by social-paediatric centres that are commonly located at paediatric hospitals. Some interviewees highlighted that families with children with behavioural problems may be less likely to attend appointments anyway and may have been persuaded to miss appointments following public messages not to visit healthcare facilities if avoidable. However, there was broad agreement that appointments that were suspended from the side of care providers were a major problem (see box 1a for illustrative statements).

### Box 1

Illustrative statements in reply to the interview question concerning patient groups treated worse during the pandemic and possible solutions

#### a on interrupted therapies and cancelled clinic appointments

*Int. 5: “Certain key people […] don’t necessarily see the parents’ suffering. I believe at the point where we from primary care [refer these patients] they have already gone a long way. […] And this is really tough, they will suffer from that for their whole lives*.*”*

*Int. 3: “Everything closed down, the social workers did no longer come out: this means, those who are not able to care for their children, they were undisturbed in not caring. The speech therapists did not work, nobody did anything. Passing time is really valuable in a child, half a year is a lot for a 3-year-old*.*”*

*Int. 2: “The problem is that their care is not just under me, but also under paediatric psychiatrists and in social-paediatric centres […] where no visits took place for a long time. And therapies, occupational therapy, speech and language therapy – there were many breaks there, where they now have to start from the beginning*.*”*

*Int. 5: “In parts, I’m disappointed. From the outside it looks like hospitals had just laid down their arms – thrown themselves on their backs like a dog and put their feet up. […] it’s exactly these children who had been waiting for an appointment for nine months […] It’s a huge fight to get somewhere anyway, and then these appointments are all cancelled*.*”*

#### b on how to increase patient trust in healthcare facilities

*Int. 4: “And the people are aware now, the hygiene precautions are understood, the practices are equipped. […] The patients don’t need to be afraid to contract something in the practices someway. Exactly this should be shown with pictures and videos, what the practices look like from the inside, how trained and equipped staff approach patients with protective equipment, that this is just illustrated - so the people see that they can feel safe in a practice, that you’re not going to catch anything there*.*”*

Interviewees were uncertain about possible solutions to this problem and expressed a high degree of acknowledgement for the difficulties in keeping up therapies and specialist clinics. However, regarding therapies there was agreement that for lack of available therapists it will not be possible to compensate for the lost time. For clinic appointments, interviewees suggested that hospitals should increase their efforts to contact families whose appointments were cancelled in order to offer alternatives with as little delay as possible.

The other concern mentioned in multiple interviews were families who were reluctant to seek medical care for fear of contracting COVID-19. This problem was felt to affect both patients with chronic conditions who missed regular follow-up appointments and patients with acute illness who were sometimes seen to present later than they usually would have. Here interviewees suggested that there should be clear and unambiguous public communications from officials that seeking medical care is safe despite the ongoing pandemic (box 1b).

### Box 2

Illustrative statements in reply to the interview question concerning worries regarding patient care during the upcoming winter

#### a on mildly symptomatic children presenting for organisational reasons

*Int. 1: “A snuffly child is presented and the mother says: ‘Normally I would’ve gone buy a pack of tissues’, now the child is not allowed to go to kindergarten and the mother is under pressure, she has to go to work and I have to confirm that the child is allowed to attend kindergarten – which I can’t*.*”*

*Int. 5: “The main problem will be frustrated parents, and that’s what I’m most afraid of personally, because I will again be their last resort and they will want a solution from me, but I don’t have one*.*”*

*Int. 3: “They’re coming in with the mission to be issued a sick note for their employer. […] They are aware that the child doesn’t need a doctor. And that’s a difference, if they present because I should issue a rag [German colloquial derogatory term for a document] or because they need a doctor*.*”*

*Int. 1: “And my worry is, how are we going to cope with this workload? -which is pointless anyway. The patient doesn’t benefit. If we had sick children, then we would need to offer 24-hour service at a pinch. But these children are not unwell, they have a cold*.*”*

#### b on the difficulties of distinguishing COVID-19 from other respiratory infections

*Int. 7: “Until now it’s still possible not to have too many contacts in the practice, because there are not that many patients, so there are no long waits. But when you think of last winter, where people needed to wait for two hours because there are too many and I’m on my own, then that’s impossible organisationally*.*”*

*Int. 6: “And we will have to pay a lot of attention that people stick to their appointments. […] As I know our parents, it will still happen time and again that the acutely sick child coughing and with a fever, will just come in and stand here. That will push us over the edge in terms of organisation*.*”*

#### c on the illness of team members and need to refer for testing

*Int. 3: “And I as a paediatrician just cannot distinguish between all these viruses […] – that means I have endless viral infections and some of it will happen to be corona. I don’t believe it will harm the children. I have gone to great lengths and made it my mission that no one from my team catches it and it worked, but I’m afraid I won’t keep that up. […] if I can’t work, that’s obviously dramatic when you’re self-employed, but also if multiple of my assistants drop out, I haven’t even considered that, but I won’t be to work*.*”*

*Int. 1: “A huge problem is when one of my staff has respiratory symptoms, we already had that three times – all colleagues from other specialties are allowed to swab their employees, but we’re only allowed to treat patients until the age of eighteen, that means I’m not allowed to swab my assistant, although we would have a [separate workspace] for example for telephone receptionist duties, and with a bit of a cold she isn’t unfit for work. But I want to separate and not let her [work with patients] unless I know she’s definitely negative. I can’t do that, I’ll have to give her leave so she can go to her GP and they will give her a sick note for three days to be safe, because only then the test result will be back – if this continues, all paediatricians will be left without staff this winter*.*”*

Concerns for the upcoming winter season universally centred on shortages of resources, mainly of staff and time which were seen as interconnected. Figure 2 shows the topics named as worrying interviewees the most regarding the upcoming winter.

**Fig. 2:**
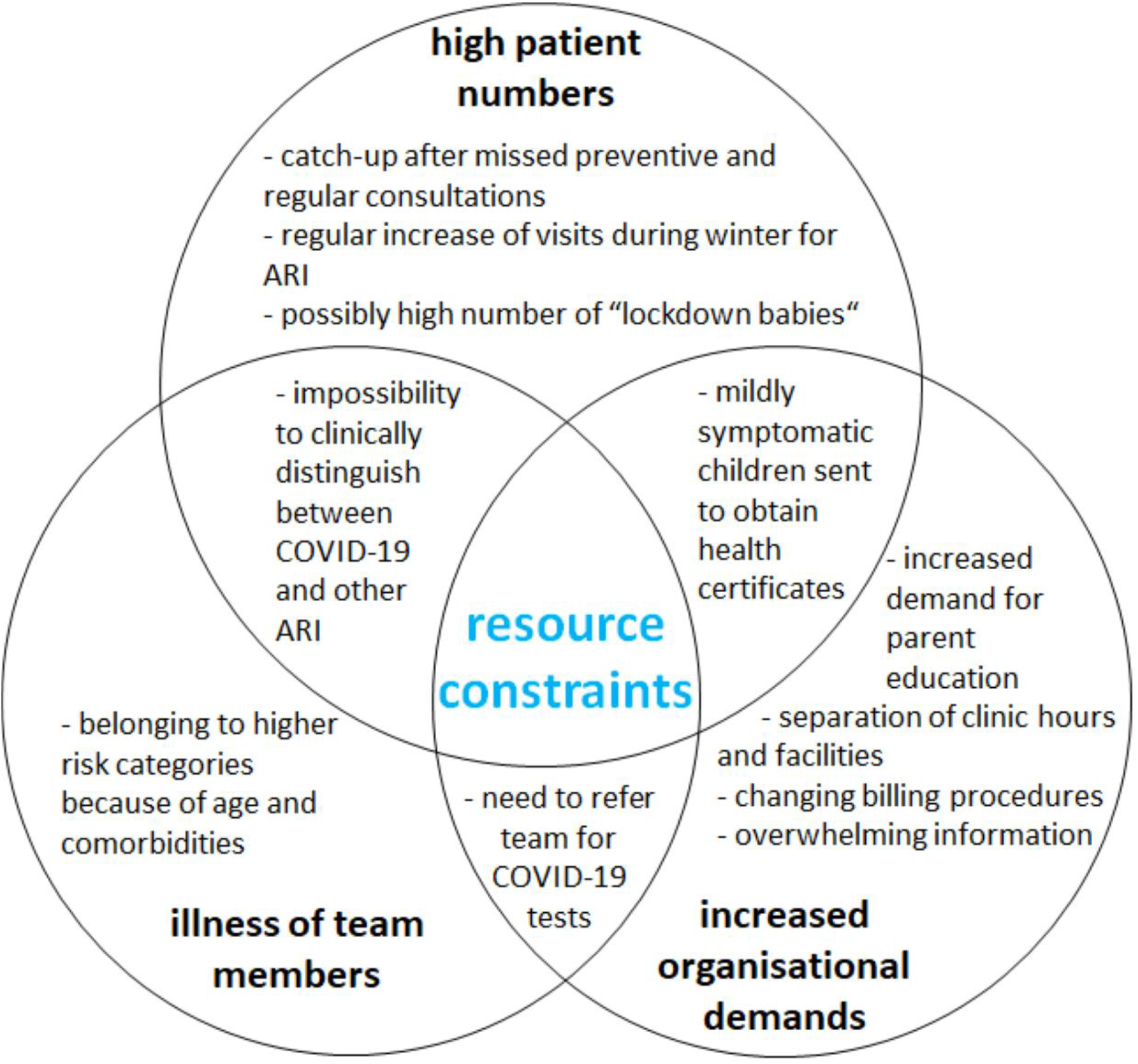
Venn diagram [25] of topics worrying primary care paediatricians most when thinking about patient care in the upcoming winter; ARI: acute respiratory infection, COVID-19: coronavirus disease 2019 – resource constraints in the intersectional areas are further illustrated by selected interviewee responses (see box 2)

Most frequently mentioned was a high number of patients expected to present with mild symptoms who require a certificate that their symptoms are not caused by SARS-CoV-2 or a sick note for their child so that they, their parents, would be able to stay home with them. This was frequently illustrated by examples from current practice in combination with the statement that this was felt to be frustrating even now where patient numbers are low, but te real concern is that it would lead to a collapse once these consultations compete with those of more severely ill patients (see box 2a for illustrative statements).

Asked for possible solutions, interviewees highlighted the need for improved and unambiguous guidelines for day-care centres and schools. They repeatedly explained that the guidance currently issued to day-care centres by the state was both hard to understand and left too many details up to interpretation, thinking that any ambiguity will lead to day-care centres understandably shifting responsibility back to providers of medical care.

Further, interviewees were consistently worried that with rising patient numbers they will need to invest increasing amounts of resources into separating possibly contagious children from others and from their staff, with the added problem that clinical criteria for potential COVID-19 patients will be unable to distinguish them from children with common respiratory infections (box 2b,c).

Multiple interviewees expressed positive views about their general preparedness for the winter season. Specifically, they highlighted that they felt sufficiently equipped with materials for infection prevention, including personal protective equipment. Also, interviewees expressed that they felt they were working in a health system showing high capacity to adapt to the current situation, including flexibility from public health authorities and health payers.

Some interviewees explained that they felt population surveillance might be helpful to have the best possible knowledge of disease activity at any time, and to better be able to adjust their level of caution. They also expressed that definite guidelines and screening questions on what patients may be at risk of being infected with SARS-CoV-2 would be helpful. However, a view expressed multiple times was that this may not be possible because of a requirement of continuous changes during the evolving pandemic.

Availability of information on COVID-19 and the evolution of the pandemic was not seen as a problem by the interviewees. Some mentioned that they felt the information offered was almost too much to handle, but generally interviewees felt they were able to extract everything necessary for their daily practice. The most commonly named sources of information were official communications and websites by the RKI, medical journals and the online members’ area offered by the paediatricians’ trade association (Berufsverband, bvkj) that allows sharing of documents and discussion among users. Multiple interviewees felt they would benefit from filtered and structured updates from a trusted source. Two interviewees mentioned the importance of informal exchange among community paediatricians for forming consensus on how to overcome problems and meet challenges, for example during continuing education meetings or informal evening meetings (that were difficult to maintain especially early in the pandemic).

## Discussion

In line with previous studies from hospital emergency departments, this survey showed that the reduction of patient consultations during the first wave of the COVID-19 pandemic was largely due to reduced unscheduled visits. Hospital emergency departments reported a similar reduction in emergency consultations [22]. Therefore, while part of this reduction may be explained by changes in care-seeking behaviour, it seems probable that visit numbers were at least partly reduced due to the generally lower incidence of acute respiratory infections in the general population [3]. A recent study showed that in Germany the proportion of children newly diagnosed with type 1 diabetes who presented with ketoacidosis was increased during the early pandemic [10]. It is therefore likely that another factor contributing to lower emergency consultation numbers may indeed be reluctance to seek care even in medically severe situations. While consultation numbers for regular developmental examinations only changed by a small degree, all surveyed paediatric practices reported that parents cancelled scheduled appointments. The observation that developmental examinations during the first year of life continued unchanged, while they were slightly reduced for older children, may reflect that narrower suggested windows for these examinations increased the willingness to keep scheduled appointments. Although completing routine vaccination schedules decreased in the United Kingdom in the current pandemic [16], seeking of routine vaccinations may still have contributed to the consistency of visits as well. Following the interviews, it seems most likely that reported cancellations are often children with chronic conditions who missed scheduled follow-up appointments. This finding is concordant with observations from previous epidemics and other settings, and a need to prepare for a higher number of follow-up visits as compensation has been stated [9,14]. Video consultations for patients with chronic conditions have been shown to be feasible across a range of settings [6,18]. These may be a good option for a group of patients reluctant to have face-to-face visits. Interruptions in regular therapies, like speech and language therapy or physiotherapy, have been highlighted as a problem across a range of countries [5,9,11].

Clinical signs are unspecific and it is clinically impossible to discern COVID-19 from other acute respiratory infections [24]. Although, the majority of care providers adopted official case definitions and used these to guide testing, this proportion was slightly lower than the proportion documented in a survey in paediatric hospital emergency departments [13]. A perception of lower applicability of case definitions for the primary care setting may have influenced this behaviour. Performance of predictive scores depends on the population they are applied to and scores developed in inpatient care may be less suitable for primary care [8]. Establishing aetiology is further complicated by the long turnaround times for SARS-CoV-2 tests [13]. There is limited data available on presenting features and the course of disease in children managed as outpatients. Multiple registries in Germany compete for hospitalised COVID-19 patients [12], but none follow-up outpatients. This is an important area for future research.

The main limitation of this study is the small scope with a limited number of surveyed and interviewed care providers in a small geographical area. Yet, the survey had a good response rate among paediatric practices representing a broad spectrum in the Düsseldorf area and the findings are likely to be transferable to other similar settings in Germany and internationally. Population management strategies for COVID-19 have been proposed, but these are not specific for Paediatrics [1]. Paediatric primary care, both in hospital emergency departments and in the community, would benefit from clear testing criteria specific for children and adolescents, that incorporate the considerations most relevant for these age groups. Important criteria that need to be taken into account are:

- lower individual risk of severe disease [7,27]
- increased mixing compared to the adult population [4,20]
- inability to quarantine on their own
- very high prevalence of non-specific respiratory symptoms during winter in young children [15,17]
- changing probability of COVID-19 or alternative diagnoses (RSV, influenza) depending on calendar month and incidence of COVID-19 in the general population

## Supporting information

Survey and consent form (German)

## Data Availability

The datasets and interview transcripts analysed for this article are available from the corresponding author on reasonable request.

## Statements

## Acknowledgements

The authors would like to thank Isabelle Hubbard for reviewing and improving the language of the manuscript and the participating health care providers for finding time to contribute to the study despite the challenges and time pressures they are facing in their daily practice.

