## Supplementary material for "Primary care during the COVID-19 pandemic – observations and challenges identified through a survey and interviews with community paediatricians: the calm before the storm": Survey and consent form (German)

UKD Universitätsklinikum Düsseldorf — hhu Heinrich Heine Universität Düsseldorf

### Fragebogen niedergelassene Kinderärzte – COVID-19-Pandemie

#### *Fragen zur Praxis*

Stadt, Stadtteil (jeweils Großraum):

O Düsseldorf, Neuss – O Leverkusen – O Mettmann, Hilden – O Ratingen, Heiligenhaus, Velbert – O Wuppertal, Solingen – O Grevenbroich, Mönchengladbach – O Duisburg, Krefeld, Moers, Oberhausen – O Andere:

Lage: O Innenstadt – O Stadtrand – O Vorstadt – O ländlich

Hat Ihre Praxis weitere Zweigstellen? O ja – O nein

Wenn ja, wo und Lage wie oben:

Wie viele Mitarbeiter hat Ihre Praxis (alle Berufsgruppen mit Patientenkontakt):

Welchem Krankenhaus weisen Sie im Regelfall zu:

oder O keine Angabe

#### *Fragen zu COVID-19-Patienten in Ihrer Praxis*

Nutzen Sie eine standardisierte Falldefinition um festzustellen, welcher Patient ein COVID-19-Verdachtsfall ist? O ja – O nein

Wer hat diese Falldefinition herausgegeben? O RKI – O lokales Gesundheitsamt – O andere:

Haben Sie Abläufe so geändert, dass Patienten mit Verdacht auf COVID-19 frühzeitig von anderen Patienten getrennt werden?

O ja, Verdachtsfälle werden an einen anderen Ort (z.B. Krankenhausnotaufnahme, Gesundheitsamt) umgeleitet

O ja, getrennte Räumlichkeiten

O ja, getrennte Sprechstundenzeiten

O nein, der Verdacht wird erst regulär während der Konsultation festgestellt

O andere Angabe und / oder Ergänzungen:

Wonach entscheiden Sie, welche Patienten Sie auf eine Infektion mit SARS-CoV-2 testen?

O nur Verdachtsfälle nach oben genannter Falldefinition – O wie zuvor, aber mit Ausnahmen (einige zusätzliche Patienten, die die Falldefinition nicht erfüllen) – O andere Kriterien (bitte führen Sie diese weiter unten aus) – O Einzelfallentscheidung

Andere Kriterien:

Können Sie sagen, wie viele Patienten Sie in den aufgeführten Wochen auf eine Infektion mit SARS-CoV-2 getestet haben (Virusnachweis, d.h. PCR-Test)?

- KW 9 (24.2. – 1.3.): oder O nicht ermittelbar
- KW 13 (23.3. – 29.3.): oder O nicht ermittelbar
- KW 17 (20.4. – 26.4.): oder O nicht ermittelbar
- KW 22 (11.5. – 17.5.): oder O nicht ermittelbar

Wie lange dauert es, bis Sie ein Testergebnis zurückerhalten?

O frühestens am gleichen Tag – O frühestens am Folgetag – O oft mehr als einen Tag später

Haben Sie bereits Patienten serologisch auf Antikörper gegen SARS-CoV-2 untersucht? O ja – O nein

Falls ja: Wie entscheiden Sie, welche Patienten sie serologisch untersuchen?

O Gleiche Kriterien wie für PCR-Test – O Andere Kriterien als für PCR-Test (bitte weiter unten aufführen) – O Einzelfallentscheidung

Andere Kriterien:

#### *Fragen zur allgemeinen Versorgung von Patienten in Ihrer Praxis seit Beginn der COVID-19-Pandemie*

Unterscheiden sich Ihre Patientenzahlen zwischen diesem und letztem Jahr, d.h. von März bis Mai 2020 im Vergleich zu den Vorjahren (März-Mai):

O Ja, mehr Patienten – O Ja, weniger Patienten – O in etwa gleich viele – O keine Angabe

Wenn ja: Können Sie dies in Prozent ausdrücken? ________________

Musste Ihre Praxis während der Pandemie zeitweise ganz oder teilweise (z.B. tageweise oder an einzelnen Standorten) schließen (nicht: ohnehin geplanter Urlaub; Mehrfachantwort möglich)

O Ja, aus eigener Initiative – O Ja, wegen eigener Krankheit – O Ja, wegen äußerer Vorgaben – O Nein – O keine Angabe

Wenn ja: Können Sie dies in Arzttagen (1 Arzt/Ärztin und 1 Tag) ausdrücken? _________________

Haben Sie bereits vereinbarte Termine mit Patienten wegen der Pandemie abgesagt?

O Ja, aus eigener Initiative – O Ja, wegen äußerer Vorgaben – O Nein – O keine Angabe

Haben Sie bereits vereinbarte Termine mit Patienten wegen der Pandemie verschoben?

O Ja, aus eigener Initiative – O Ja, wegen äußerer Vorgaben – O Nein – O keine Angabe

Haben Sie bereits vereinbarte Termine mit Patienten telefonisch durchgeführt?

O Ja, aus eigener Initiative – O Ja, wegen äußerer Vorgaben – O Nein – O keine Angabe

Wurden vereinbarte Termine von Eltern wegen der Pandemie abgesagt oder verschoben?

O ja – O nein – O keine Angabe

Haben Sie Termine für Vorsorgeuntersuchungen (U, J) abgesagt?

O Ja, aus eigener Initiative – O Ja, wegen äußerer Vorgaben – O Nein – O keine Angabe

Bitte tragen Sie Ihre Konsultationszahlen für einzelne Anlässe und Kalenderwochen im Vergleich 2019 – 2020 in die Tabelle auf der nächsten Seite ein

Gibt es bestimmte Patientengruppen, die Ihrer Meinung nach während der Pandemie in Ihrer Praxis <u>schlechter</u> als bisher versorgt werden? O ja – O nein – O unsicher – O keine Angabe

Wenn ja, welche:

Gibt es bestimmte Patientengruppen, die Ihrer Meinung nach während der Pandemie in Ihrer Praxis <u>besser</u> als bisher versorgt werden? O ja – O nein – O unsicher – O keine Angabe

Wenn ja, welche:

*Stattgefundene Konsultationen:*

| 2020 | | | | 2019 | | | |
|---|---|---|---|---|---|---|---|
| KW | U1 – U6 | U7 – J2 | Notfall | KW | U1 – U6 | U7 – J2 | Notfall |
| 9 (24.2. – 1.3.) | | | | 9 (25.2. – 3.3.) | | | |
| 13 (23.3. – 29.3.) | | | | 13 (25.3. – 31.3.) | | | |
| 17 (20.4. – 26.4.) | | | | 19 (6.5. – 12.5.) | | | |
| 22 (25.5. – 31.5.) | | | | 21 (20. – 26.5.) | | | |

oder

O diese Zahlen kann ich nicht ermitteln (für 2020)

O diese Zahlen kann ich nicht ermitteln (für 2019)

*Abgesagte und verschobene Konsultationen:*

| **2020** KW | von Praxisseite abgesagt | | von Praxisseite verschoben | | von Patientenseite abgesagt/verschoben | |
|---|---|---|---|---|---|---|
| | U1 – U6 | U7 – J2 | U1 – U6 | U7 – J2 | U1 – U6 | U7 – J2 |
| 9 (24.2. – 1.3.) | | | | | | |
| 13 (23.3. – 29.3.) | | | | | | |
| 17 (20.4. – 26.4.) | | | | | | |
| 22 (25.5. – 31.5.) | | | | | | |

oder

O diese Zahlen kann ich nicht ermitteln

Vielen Dank, dass Sie sich die Zeit nehmen, diesen Fragebogen auszufüllen! Bitte beachten Sie die Datenschutzhinweise und die Fragen zu weiteren Studien auf den kommenden Seiten!

UKD Universitätsklinikum Düsseldorf

hhu Heinrich Heine Universität Düsseldorf

#### Aufklärung und Einwilligung zur Datenverarbeitung

Wir versichern Ihnen, dass die Forschungsdaten nur dazu verwendet werden, den wissenschaftlichen Fragen bezüglich der Arbeitssituation in Praxen in Zeiten des neuen Coronavirus nachzugehen. Die Verwendung der Daten zu anderen Zwecken ist ausgeschlossen und aufgrund der Datenschutzverordnung nicht zulässig. Selbstverständlich ist die Teilnahme freiwillig. Ihnen entstehen keine Nachteile, wenn Sie sich entscheiden nicht an der Studie teilzunehmen.

Die Studie folgt streng den Bestimmungen des Datenschutzes. Einwilligungserklärung und Fragebögen werden getrennt voneinander gesammelt. Dieses Blatt wird nach Eingang von den anderen Seiten des Fragebogens getrennt und separat verwahrt. Eine Zuordnung der Angaben auf den anderen Seiten zu Ihnen oder Ihrer Praxis ist dann nicht mehr eindeutig möglich. Ohnehin wird Ihr Name oder der Name Ihrer Praxis bei der Auswertung der Fragebögen und einer möglichen Darstellung der Ergebnisse nicht genannt. Die Darstellung der Ergebnisse erfolgt aggregiert (d.h. als Ergebnisse der gesamten Gruppe oder sinnvoller Teilgruppen) und jede Gruppe enthält die Ergebnisse von mindestens mehr als einer Praxis. Die anonymisierten Forschungsdaten werden für 10 Jahre gespeichert. Die Weitergabe der anonymisierten Forschungsdaten ist nur an bekannte Personen/Institutionen gemäß Datenschutz-Grundverordnung (DSGVO) möglich.

Zugriff auf die Studiendatenbank haben nur befugte Mitarbeiter/innen, die alle auf das Berufs- und Datengeheimnis verpflichtet sind. Die Fragebögen werden im Studienzentrum gesammelt und verschlossen aufbewahrt.

**Herr Dr. Malte Kohns Vasconcelos, als Studienleiter, ist für die Datenverarbeitung und Auswertung im Studienzentrum verantwortlich. Bei Beschwerden können Sie sich an Herrn Malte Kohns Vasconcelos oder als Beschwerdeinstanz an die Stabsstelle Datenschutz oder an die Aufsichtsbehörde (siehe Rückseite) wenden. Rechtsgrundlage für die Verarbeitung der erhobenen Daten ist die Datenschutz-Grundverordnung in Verbindung mit dem Bundes- und Landesdatenschutzgesetz.**

Sie dürfen Ihre Zustimmung zu dieser Studie zu jedem Zeitpunkt und ohne Begründung widerrufen. Ihnen entstehen dadurch keinerlei Nachteile. Bei Ihrem Widerruf werden alle Daten, die noch nicht verarbeitet worden sind, gelöscht. Bereits anonymisierte Daten können nicht gelöscht werden, sind aber nicht mehr rückverfolgbar.

**Einwilligung:**

Ich habe das Anschreiben sowie dieses Informationsblatt zur vorliegenden Studie gelesen und wurde damit über die Ziele und den Ablauf der Studie, sowie über den Umgang mit den in der Studie erhobenen Datenaufgeklärt.

Nach dieser Aufklärung willige ich in die Verarbeitung der von mir eingegebenen Daten ein.

**Ort und Datum:**

**Ausfüllende Person:**

**Email-Adresse:**

**Unterschrift:**

**Verantwortlicher für die Datenverarbeitung:**
Dr. med. Malte Kohns Vasconcelos
Institut für Medizinische Mikrobiologie und Krankenhaushygiene, Geb. 22.21
Universitätsklinikum Düsseldorf, Moorenstr. 5, 40225 Düsseldorf
Fon: +49 (0) 211 81 14886


UKD Universitätsklinikum Düsseldorf — hhu Heinrich Heine Universität Düsseldorf

**Beschwerdeinstanz:**
Stabsstelle Datenschutz
Datenschutzbeauftragte UKD
Moorenstraße 5
40225 Düsseldorf
Fon: +49 (0) 211 81 00 (Zentrale)


**Aufsichtsbehörde** – im Falle einer rechtswidrigen Datenverarbeitung haben Sie das Recht, sich hier zu beschweren:
Landesbeauftragte für Datenschutz und Informationsfreiheit Nordrhein-Westfalen
Postfach 20 04 44, 40102 Düsseldorf


#### *Fragen zu weiteren Studien*

Wären Sie prinzipiell bereit, an einem telefonischen Interview zur Versorgung von Patienten in Ihrer Praxis während der Pandemie teilzunehmen? O ja – O nein

Hätten Sie ein prinzipielles Interesse, mit den Mitarbeitern Ihrer Praxis an einer Studie zur Seroprävalenz von SARS-CoV-2-Antikörpern bei Mitarbeitern in der pädiatrischen Primärversorgung teilzunehmen?

O ja, auch ohne, dass wir personalisierte Befunde personalisierte Befunde erhalten und damit davon keinen direkten Nutzen haben

O ja, aber nur, wenn für die Untersuchungsergebnisse personalisierte Befunde für meine Mitarbeiter erstellt werden

O nein

Hätten Sie Interesse, klinische Verlaufsdaten der COVID-19-Patienten in Ihrer Praxis in einer zentralen Datenbank zu sammeln?

O ja, ich würde Patienten dafür auch aufklären und gegebenenfalls zu einem festgelegten Zeitpunkt nach Diagnosestellung aktiv nach Ihrem Wohlergehen befragen

O ja, aber ich würde nur anonymisiert Daten erheben und nur, wenn dazu keine Patientenaufklärung erforderlich ist

O nein, kein Interesse

Ich selbst oder Mitarbeiter meiner Praxis haben Interesse, an der Auswertung und Veröffentlichung einer dieser Untersuchungen im Rahmen der COVID-19 mitzuwirken. O Ja
